# Nutritional status, clinical burden, and healthcare utilization among pediatric outpatients with congenital heart disease: A retrospective cross-sectional study from Indonesia

**DOI:** 10.64898/2026.05.23.26353925

**Authors:** Putri Amelia, Lourents Christian Dika Sahertian, Rizky Adriansyah, Johnson Kannady

**Author notes:** Corresponding Author: Name: Putri Amelia Current Institution: Universitas Sumatera Utara.

## Abstract

Congenital heart disease contributes substantially to chronic morbidity, growth impairment, and repeated healthcare utilization among children. Evidence regarding nutritional burden and outpatient healthcare patterns among pediatric patients with congenital heart disease in Indonesia remains limited. This study aimed to evaluate clinical characteristics, nutritional status, healthcare utilization, and factors associated with malnutrition among pediatric outpatients with congenital heart disease at Adam Malik General Hospital, Indonesia. A retrospective observational study was conducted using medical records of pediatric outpatients treated between January and December 2024. Demographic characteristics, cardiac diagnoses, nutritional status, complications, and outpatient visit history were analyzed. Logistic regression analysis was performed to identify factors associated with malnutrition. A total of 606 pediatric outpatients were included. Non cyanotic congenital heart disease predominated the cohort, with ventricular septal defect representing the most common diagnosis followed by patent ductus arteriosus and atrial septal defect. Nearly half of all patients demonstrated underweight or severe underweight nutritional status, while pulmonary hypertension emerged as the most frequent complication. Younger pediatric age groups and higher cumulative clinical burden independently increased the odds of malnutrition. Children with congenital heart disease at this tertiary referral center carried a substantial nutritional and clinical burden. Early nutritional surveillance and integrated long term outpatient management may improve growth outcomes and reduce chronic disease burden in resource limited settings.

## INTRODUCTION

Congenital heart disease (CHD) represents the most common congenital anomaly in children and remains a major contributor to pediatric morbidity and mortality worldwide. Structural abnormalities of the heart alter normal hemodynamics from birth and often lead to chronic clinical complications throughout childhood. Improvements in neonatal care, echocardiographic evaluation, interventional cardiology, and pediatric cardiac surgery have increased survival rates among children with CHD. Despite these advances, many patients in low and middle income countries still face delayed diagnosis, limited access to specialized cardiac services, and prolonged exposure to untreated hemodynamic disturbances. These conditions frequently impair growth, nutritional status, and overall quality of life.[1–3]

Acyanotic lesions such as ventricular septal defect (VSD), atrial septal defect (ASD), and patent ductus arteriosus (PDA) account for the majority of pediatric CHD cases worldwide. Persistent left to right shunting increases pulmonary blood flow and cardiac workload which may progress to pulmonary hypertension, recurrent respiratory tract infections, and heart failure. Cyanotic lesions including tetralogy of Fallot (TOF) and transposition of the great arteries (TGA) produce chronic hypoxemia that further disrupts systemic oxygen delivery and tissue metabolism. Ongoing circulatory abnormalities increase energy expenditure while reducing feeding tolerance and nutrient utilization. Children with CHD therefore face a substantial risk of growth failure and malnutrition during critical periods of physical development.[1,4–6]

Malnutrition remains one of the most important comorbid conditions among pediatric patients with CHD because nutritional impairment directly affects immune function, neurodevelopment, surgical readiness, and long term clinical outcomes. Children with severe undernutrition often require more frequent hospital visits and prolonged medical follow up because poor nutritional reserve worsens susceptibility to infections and delays clinical recovery. Previous studies have shown that younger children with CHD experience higher rates of nutritional problems due to rapid growth demands combined with increased metabolic stress from chronic cardiac dysfunction. The interaction between cardiac disease severity, feeding difficulty, recurrent symptoms, and healthcare utilization creates a complex clinical burden that extends beyond the primary structural defect itself.[4–8]

Several studies from Indonesia have described the demographic and clinical characteristics of pediatric CHD populations in tertiary referral hospitals. Most reports consistently identify VSD, ASD, and PDA as the predominant lesions among pediatric patients. Earlier investigations also documented high rates of respiratory complications and nutritional impairment among affected children. However, many available studies remain limited to descriptive profiling without evaluating factors associated with malnutrition or healthcare burden in outpatient settings. Evidence from Indonesia regarding the relationship between nutritional status, clinical burden, and healthcare utilization among pediatric CHD outpatients also remains limited.

Adam Malik General Hospital serves as a major tertiary referral center in North Sumatra and manages a large number of pediatric cardiology cases from various regions in western Indonesia. Understanding the demographic distribution, nutritional characteristics, clinical burden, and healthcare utilization patterns among pediatric CHD outpatients may provide important insight into disease burden and long term management challenges in resource limited settings. This study therefore aimed to evaluate the clinical profile of pediatric CHD outpatients at Adam Malik General Hospital in 2024 with emphasis on nutritional burden, healthcare utilization, and factors associated with malnutrition.

## METHODS

### Study design and setting

This study used a retrospective observational design based on secondary data obtained from medical records of pediatric outpatients with congenital heart disease at Adam Malik General Hospital, Medan, North Sumatra, Indonesia. Adam Malik General Hospital functions as a tertiary referral center for pediatric cardiology services in western Indonesia and receives referrals from multiple districts across North Sumatra and surrounding provinces. Data collection covered outpatient visits recorded between January 1 and December 31, 2024.

### Study population and sampling

The study population consisted of all pediatric patients diagnosed with congenital heart disease who attended the pediatric outpatient clinic during the study period. A total sampling approach was applied to include all eligible patients who fulfilled the study criteria.

Inclusion criteria consisted of pediatric patients aged 0 to 18 years with a confirmed diagnosis of congenital heart disease documented through echocardiographic evaluation in the medical record. Patients were included only when demographic information, congenital heart disease classification, nutritional assessment, and outpatient visit history were available.

Patients with incomplete core demographic data or unclear congenital heart disease diagnosis were excluded from the analysis.

### Variables and operational definitions

The primary outcome of this study was malnutrition status. Nutritional status classification was based on pediatric anthropometric assessment routinely applied in the hospital using WHO growth standards and age appropriate weight for age evaluation documented in the medical record. Normal nutritional status, underweight, and severe underweight categories were classified according to standardized pediatric nutritional assessment used in routine clinical practice. Patients categorized as underweight or severe underweight were subsequently grouped into the malnutrition category for regression analysis.

Congenital heart disease was categorized into cyanotic and non cyanotic physiology. Diagnoses included atrial septal defect, ventricular septal defect, patent ductus arteriosus, pulmonary stenosis, coarctation of the aorta, tetralogy of Fallot, and transposition of the great arteries.

Demographic variables included sex and age group classification. Age categories consisted of neonates, infants, young toddlers, toddlers, preschool children, school aged children, and adolescents. Because of sparse observations in several extreme age categories, adolescent and neonatal groups were combined during the primary multivariable regression analysis to improve model stability.

Clinical variables included dyspnea, poor feeding, easy fatigability, cyanosis, poor weight gain, pulmonary hypertension, respiratory tract infection, heart failure, failure to thrive, and Eisenmenger syndrome. Healthcare utilization was assessed using the number of outpatient visits documented during the study year.

A cumulative burden score was developed to represent overall clinical burden. The score was calculated from the total number of documented symptoms and complications identified in each patient. Higher burden scores indicated greater cumulative clinical burden.

### Data collection procedure

Medical records were reviewed systematically using hospital registration databases and outpatient cardiology records. Eligible patients were identified through diagnostic coding and echocardiographic documentation. Relevant demographic, clinical, nutritional, and healthcare utilization variables were subsequently extracted and entered into a structured data collection form. Data cleaning and verification were performed before statistical analysis to reduce entry inconsistencies and duplicate records.

### Missing data handling

Data completeness was evaluated for all study variables before analysis. Demographic variables, congenital heart disease classification, nutritional status, and outpatient visit history demonstrated complete data availability. Symptom and complication variables showed substantial missingness because retrospective clinical documentation varied between visits and treating physicians. Detailed completeness assessment and proportions of missing data across study variables are summarized in S1 Table.

Missing clinical documentation was considered likely related to inconsistent recording practices rather than true biological absence of symptoms or complications. No imputation procedures were performed because of the retrospective nature of the dataset and the non random pattern of missingness. Complete case analysis was therefore applied for regression modeling.

### Statistical analysis

Statistical analysis was performed using IBM SPSS Statistics and R statistical software. Categorical variables were presented as frequencies and percentages, whereas continuous variables were summarized using mean, standard deviation, median, minimum value, and maximum value according to data distribution.

Univariate analysis was conducted to describe demographic characteristics, congenital heart disease subtype distribution, nutritional status, clinical manifestations, complications, and healthcare utilization patterns among pediatric outpatients.

Associations between clinical variables and malnutrition were evaluated using logistic regression analysis. Univariable logistic regression was initially performed to estimate crude odds ratios and identify candidate variables associated with malnutrition. Variables considered clinically relevant or demonstrating statistical association were subsequently entered into the multivariable logistic regression model to obtain adjusted odds ratios with 95% confidence intervals.

The primary multivariable model included congenital heart disease physiology, sex, age group, and burden score as covariates. Odds ratios greater than one indicated increased odds of malnutrition. Statistical significance was defined using a two sided p value below 0.05.

Sensitivity analysis was performed using the original non collapsed age group classification to evaluate robustness of the regression findings. Multicollinearity diagnostics were assessed using generalized variance inflation factor values. Model performance was evaluated using Akaike information criterion, Bayesian information criterion, Tjur’s R², root mean square error, logarithmic score, and proportion correctly predicted.

### Ethical consideration

The Health Research Ethics Committee of the Faculty of Medicine Universitas Sumatera Utara approved this study with ethical clearance number 653/KEPK/USU/2025. Permission to access medical records was obtained from Adam Malik General Hospital before data collection. Patient confidentiality was maintained throughout the study process by anonymizing all identifiable information before statistical analysis.

## RESULTS

### Baseline characteristics of the study population

A total of 606 pediatric outpatients with congenital heart disease fulfilled the eligibility criteria and were included in the final analysis (Table 1). Female patients slightly outnumbered males with 319 cases (52.6%), whereas male patients accounted for 287 cases (47.4%). The largest age category consisted of young toddlers with 166 patients (27.4%), followed by school aged children with 138 patients (22.8%) and infants with 111 patients (18.3%). Preschool children represented the smallest subgroup with 28 patients (4.6%).

**Table 1.**
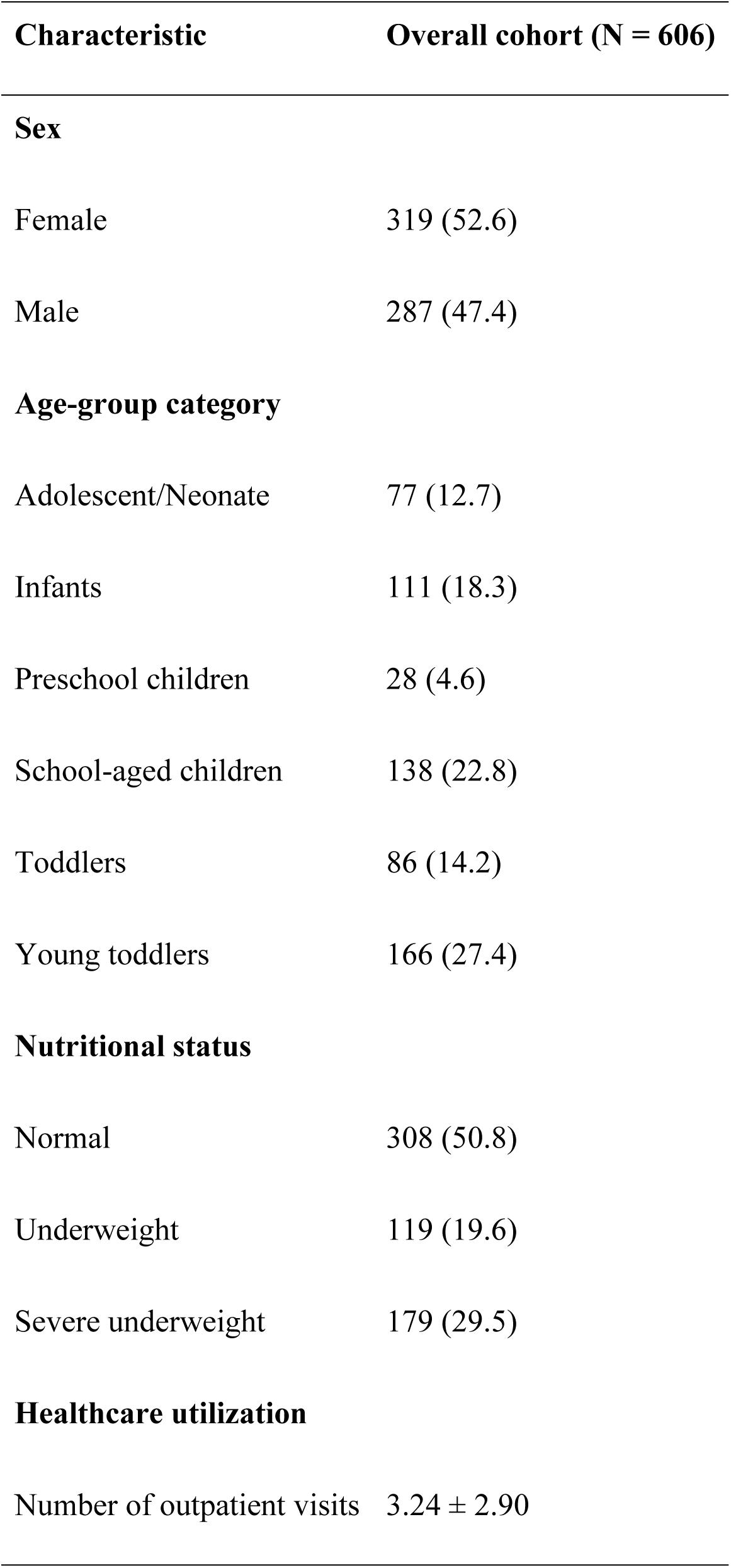
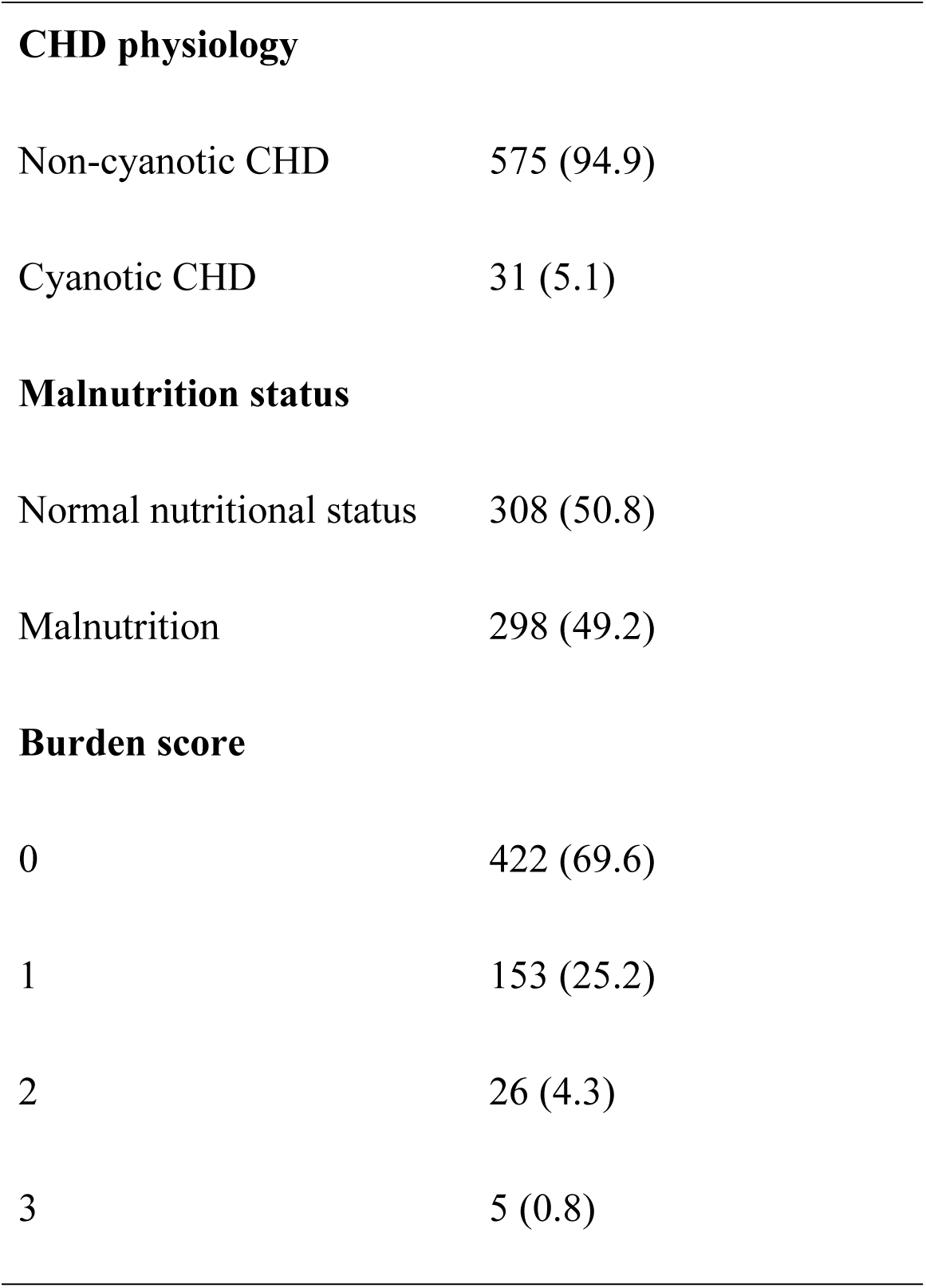
Baseline Characteristics of Pediatric Congenital Heart Disease Outpatients.

Non cyanotic congenital heart disease dominated the cohort and accounted for 575 patients (94.9%), while cyanotic lesions were identified in 31 patients (5.1%). Nutritional assessment demonstrated that 308 patients (50.8%) had normal nutritional status. Underweight and severe underweight status were identified in 119 patients (19.6%) and 179 patients (29.5%), respectively. Overall, malnutrition affected 298 patients (49.2%) within the study population.

The mean number of outpatient visits during 2024 was 3.24 ± 2.90 visits per patient. Most patients demonstrated low cumulative clinical burden scores. A burden score of 0 was identified in 422 patients (69.6%), whereas only five patients (0.8%) demonstrated the highest observed burden score of 3.

### Distribution of congenital heart disease subtypes

Ventricular septal defect represented the most frequently documented congenital heart disease subtype and was identified in 259 patients (33.5%). Patent ductus arteriosus and atrial septal defect followed closely with 218 cases (28.2%) and 217 cases (28.0%), respectively. Pulmonary stenosis accounted for 44 patients (5.7%). Cyanotic lesions occurred less frequently and included transposition of the great arteries in 18 patients (2.3%) and tetralogy of Fallot in 13 patients (1.7%). Coarctation of the aorta represented the rarest lesion with only five documented cases (0.7%)(Table 2).

**Table 2.**
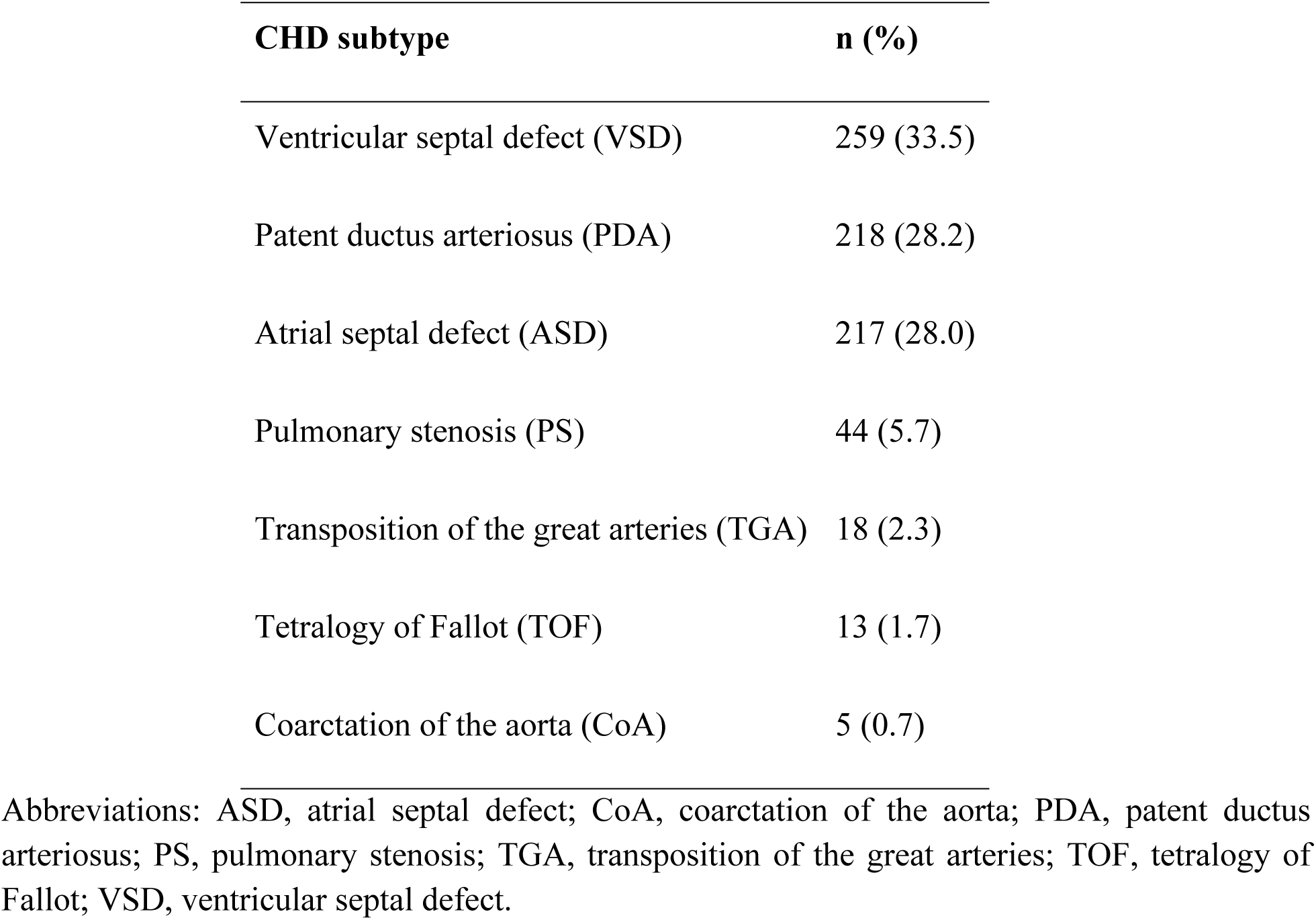
Distribution of Congenital Heart Disease Subtypes.

| CHD subtype | n (%) |
| --- | --- |
| Ventricular septal defect (VSD) | 259 (33.5) |
| Patent ductus arteriosus (PDA) | 218 (28.2) |
| Atrial septal defect (ASD) | 217 (28.0) |
| Pulmonary stenosis (PS) | 44 (5.7) |
| Transposition of the great arteries (TGA) | 18 (2.3) |
| Tetralogy of Fallot (TOF) | 13 (1.7) |
| Coarctation of the aorta (CoA) | 5 (0.7) |

Abbreviations: ASD, atrial septal defect; CoA, coarctation of the aorta; PDA, patent ductus arteriosus; PS, pulmonary stenosis; TGA, transposition of the great arteries; TOF, tetralogy of Fallot; VSD, ventricular septal defect.

### Clinical manifestations and complications

Clinical symptoms and complications varied across the cohort (Table 3). Pulmonary hypertension emerged as the most frequently documented complication and affected 115 patients (19.0%). Respiratory tract infection occurred in 56 patients (9.2%), while heart failure was identified in 46 patients (7.6%). Eisenmenger syndrome remained uncommon and was observed in only three patients (0.5%).

**Table 3.** Distribution of clinical complications among pediatric congenital heart disease outpatients.

| Complication | n (%) |
| --- | --- |
| Pulmonary hypertension | 115 (19.0) |
| Respiratory tract infection | 56 (9.2) |
| Heart failure | 46 (7.6) |
| Eisenmenger syndrome | 3 (0.5) |

Growth related manifestations also appeared frequently within the outpatient population. Poor weight gain, feeding difficulty, dyspnea, and easy fatigability were commonly documented among patients with repeated healthcare utilization and higher clinical burden scores.

### Healthcare utilization patterns

Outpatient healthcare utilization demonstrated substantial variability throughout the study period. The cumulative number of outpatient visits reached 2,065 visits during 2024. Monthly visit frequency ranged from 137 visits in April to 201 visits in July. Higher outpatient volume also appeared during October and November with 198 and 195 visits, respectively.

Evaluation of registration frequency per patient demonstrated that most patients attended outpatient services only once during the study year. Single visit registration was identified in 238 patients (39.3%). Two visits and three visits occurred in 83 patients (13.7%) and 86 patients (14.2%), respectively. A smaller subgroup demonstrated substantially higher healthcare utilization, with several patients requiring more than 10 outpatient visits within one year. The maximum recorded frequency reached 20 visits.

Patients with severe underweight status demonstrated broader healthcare utilization distributions and higher outpatient visit frequencies compared with patients who had normal nutritional status. Violin boxplot analysis also demonstrated wider variability of visit burden among malnourished patients. Detailed healthcare utilization patterns according to nutritional status are illustrated in Fig 1.

**Fig 1.**
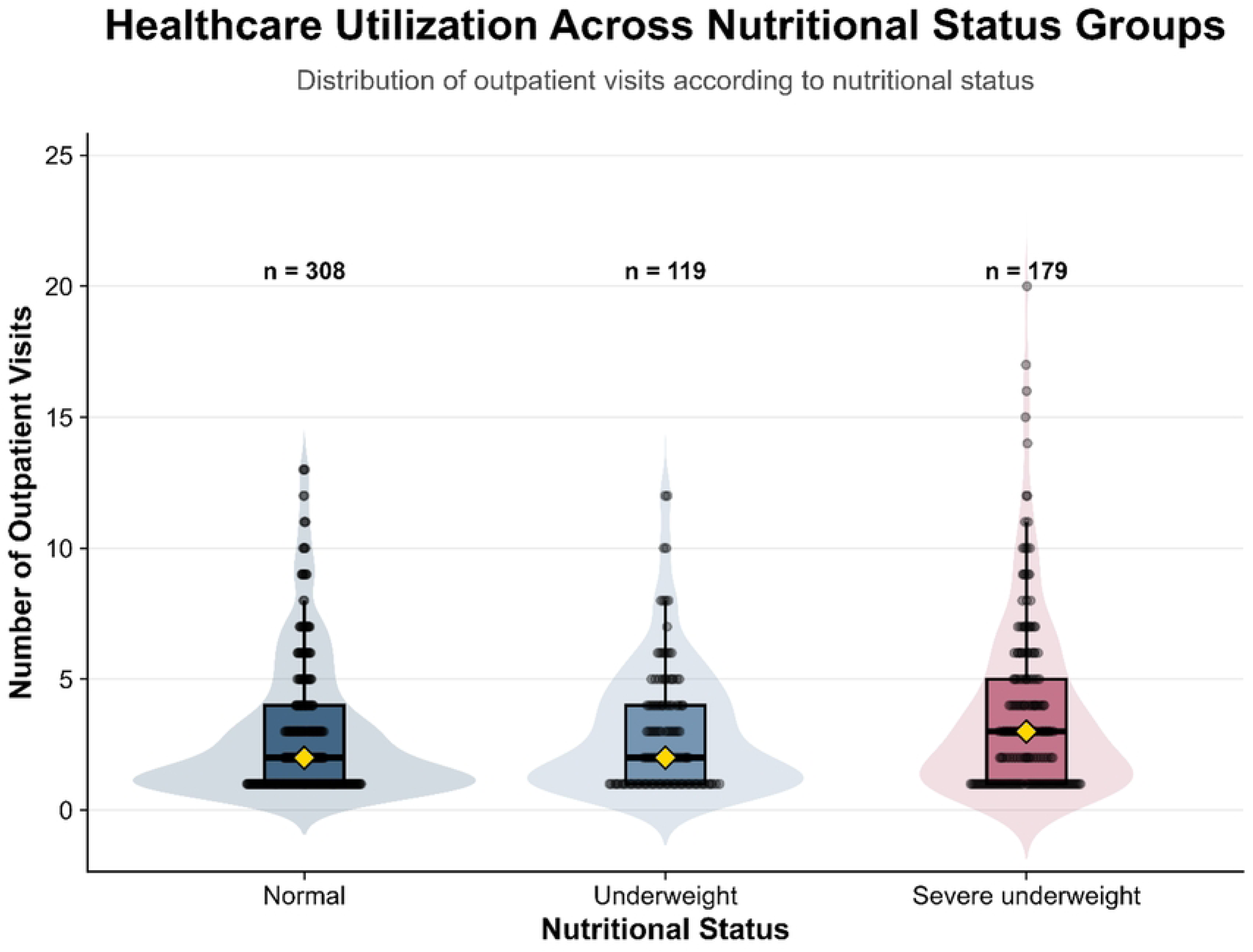
Healthcare utilization across nutritional status groups among pediatric congenital heart disease outpatients.

Violin-boxplot distribution demonstrating outpatient healthcare utilization according to nutritional status among pediatric patients with congenital heart disease. The violin component represents distribution density, whereas embedded boxplots display the median and interquartile range. Individual observations are overlaid as jittered points, and yellow diamonds indicate group medians.

### Nutritional status across congenital heart disease subtypes

Nutritional impairment appeared across both cyanotic and non cyanotic congenital heart disease groups. More than half of patients with tetralogy of Fallot and transposition of the great arteries demonstrated underweight or severe underweight status. Among non cyanotic lesions, ventricular septal defect showed the highest nutritional burden. Severe underweight and underweight collectively affected more than half of patients with ventricular septal defect.

Patients with atrial septal defect and patent ductus arteriosus more frequently demonstrated normal nutritional status compared with other lesions. Nevertheless, substantial proportions of nutritional impairment still remained evident within these groups. Coarctation of the aorta demonstrated the most severe nutritional profile because all affected patients exhibited malnutrition, although the subgroup size remained very small.

### Factors associated with malnutrition

Univariable logistic regression analysis demonstrated that high healthcare utilization, pulmonary hypertension, poor feeding, and higher burden score were associated with increased odds of malnutrition. Poor feeding demonstrated the strongest crude association and produced nearly fourfold higher odds of malnutrition compared with patients without feeding difficulties. Detailed univariable logistic regression findings are presented in S2 Table.

Multivariable logistic regression analysis demonstrated that younger pediatric age groups exhibited consistently higher odds of malnutrition compared with the adolescent and neonatal reference category (Table 4). Young toddlers demonstrated approximately 2.7 fold increased odds of malnutrition with adjusted odds ratio 2.68 (95% CI 1.52–4.74; p < 0.001). Toddlers also demonstrated significantly increased odds with adjusted odds ratio 2.28 (95% CI 1.21–4.34; p = 0.012). School aged children and infants showed elevated odds ratios, although confidence intervals crossed the null value. Distribution of nutritional status across pediatric age groups is illustrated in Fig 2.

**Fig 2.**
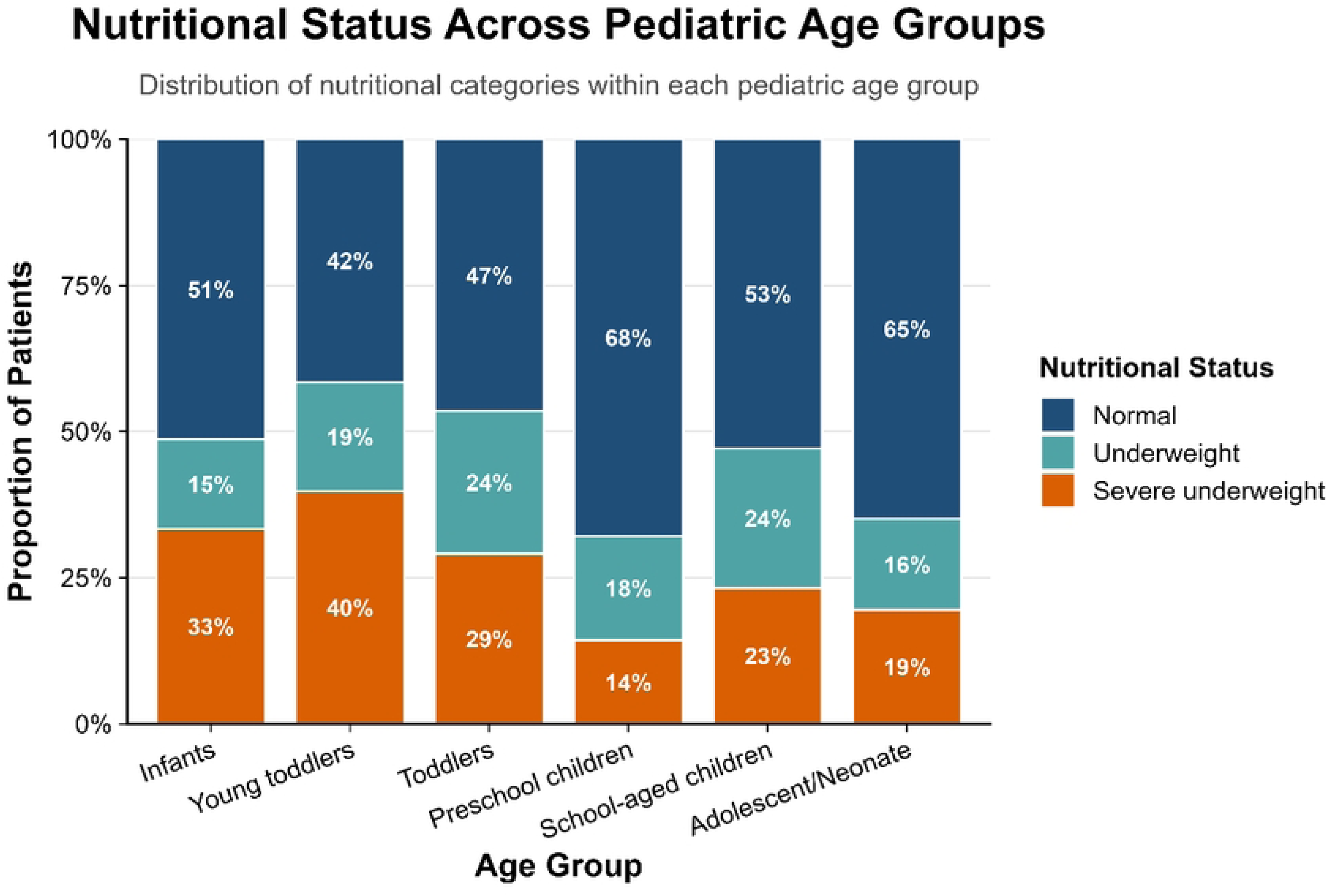
Distribution of nutritional status across pediatric age groups among patients with congenital heart disease.

**Table 4.** Multivariable Logistic Regression Analysis of Factors Associated With Malnutrition.

| Variable | Adjusted OR | 95% CI | p-value |
| --- | --- | --- | --- |
| Cyanotic CHD | 1.50 | 0.70–3.20 | 0.296 |
| Male sex | 1.10 | 0.79–1.52 | 0.579 |
| **Age group** |  |  |  |
| Infants | 1.66 | 0.91–3.05 | 0.103 |
| Preschool children | 0.83 | 0.33–2.11 | 0.694 |
| School-aged children | 1.74 | 0.98–3.12 | 0.062 |
| Toddlers | 2.28 | 1.21–4.34 | 0.012 |
| Young toddlers | 2.68 | 1.52–4.74 | <0.001 |
| Burden score | 1.50 | 1.14–1.99 | 0.004 |

Younger age categories demonstrated a higher proportion of undernutrition compared with older pediatric groups. Age-group classification followed pediatric clinical categorization routinely applied in the study institution and consisted of neonates (0–28 days), infants (1–12 months), young toddlers (13–24 months), toddlers (25–59 months), preschool children (5–6 years), school-aged children (7–12 years), and adolescents (>12 years).

Higher burden score independently remained associated with malnutrition after adjustment for covariates. Each incremental increase in burden score increased the odds of malnutrition by approximately 50% with adjusted odds ratio 1.50 (95% CI 1.14–1.99; p = 0.004).

Cyanotic congenital heart disease and male sex did not demonstrate statistically significant associations with malnutrition in the final model. Cyanotic physiology produced adjusted odds ratio 1.50 (95% CI 0.70–3.20; p = 0.296), whereas male sex produced adjusted odds ratio 1.10 (95% CI 0.79–1.52; p = 0.579). Adjusted odds ratios and confidence intervals derived from the final multivariable logistic regression model are visualized in Fig 3.

**Fig 3.**
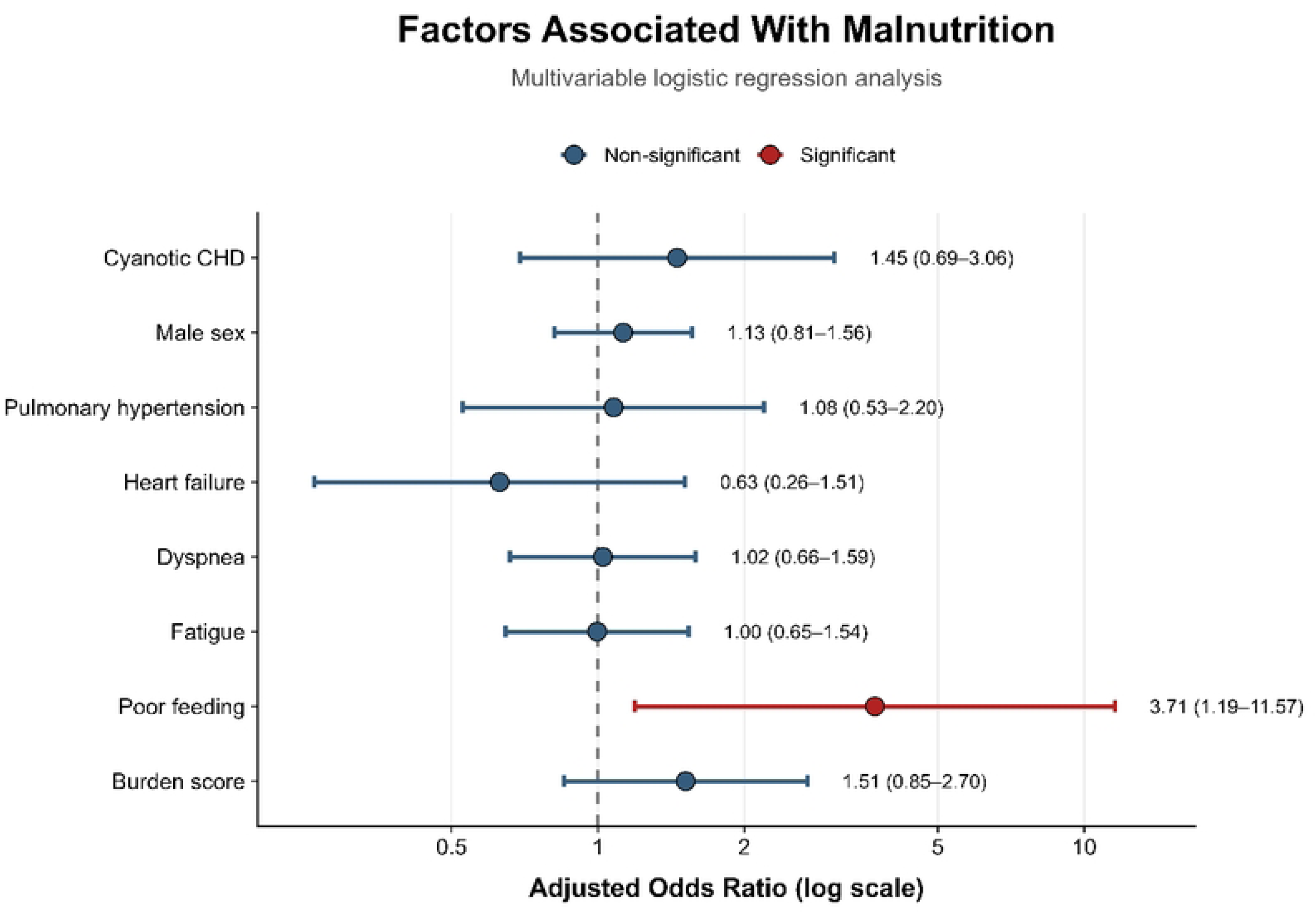
Factors associated with malnutrition among pediatric congenital heart disease outpatients.

Forest plot showing adjusted odds ratios and 95% confidence intervals derived from the multivariable logistic regression model evaluating factors associated with malnutrition among pediatric congenital heart disease outpatients.

### Sensitivity analysis and model performance

Sensitivity analysis using the original non collapsed age group classification demonstrated findings consistent with the primary model. Detailed sensitivity analysis results are presented in S3 Table. Young toddlers and toddlers remained significantly associated with increased malnutrition risk, while healthcare utilization also maintained statistical significance. Neonatal estimates could not be reliably calculated because of sparse observations and quasi complete separation within the subgroup.

Multicollinearity assessment demonstrated acceptable generalized variance inflation factor values across included covariates. No evidence of clinically significant multicollinearity was identified within the final regression model. Detailed multicollinearity diagnostics are presented in S4 Table.

Overall model performance demonstrated acceptable calibration and classification capability for exploratory clinical association analysis. Detailed model performance metrics are summarized in S5 Table. The final multivariable model produced Akaike information criterion 828.9 and Tjur’s R² 0.062. Calibration analysis demonstrated reasonable agreement between predicted and observed probabilities of malnutrition across the range of estimated risk values. Calibration performance of the final multivariable model is illustrated in S1 Fig.

## DISCUSSION

This study demonstrated that non cyanotic congenital heart disease dominated the outpatient pediatric population at Adam Malik General Hospital, with ventricular septal defect, patent ductus arteriosus, and atrial septal defect representing the most frequently identified lesions. Nearly half of all patients experienced malnutrition, while pulmonary hypertension emerged as the most commonly documented complication. Younger pediatric age groups and higher cumulative clinical burden demonstrated significant associations with malnutrition. Increased healthcare utilization also appeared more common among malnourished children, suggesting that nutritional impairment may coexist with greater disease complexity and prolonged clinical monitoring requirements.

The predominance of ventricular septal defect, patent ductus arteriosus, and atrial septal defect aligns with the global epidemiological pattern of congenital heart disease reported in both developed and developing countries. Left to right shunt lesions account for the majority of pediatric congenital heart disease because these defects occur more frequently during cardiac embryogenesis and often allow survival into childhood without immediate life threatening cyanosis. Similar distributions have been reported by previous Indonesian and international studies that consistently identified ventricular septal defect as the most common lesion among pediatric cardiology populations.[1,2,9,10]

Young toddlers represented the largest age group within the present cohort. This distribution likely reflects the period when persistent symptoms, growth disturbances, recurrent respiratory infections, or delayed developmental progress become more apparent to caregivers and healthcare providers. Children within this age range also undergo rapid somatic growth and increased nutritional demands, making underlying hemodynamic abnormalities more clinically visible. School aged children formed the second largest subgroup and may reflect long term outpatient follow up among survivors with stable congenital lesions requiring periodic cardiac evaluation. Earlier studies from tertiary referral centers have reported comparable age distributions with continued healthcare utilization extending beyond infancy.[9,11]

Nearly half of the study population demonstrated underweight or severe underweight nutritional status. This finding reinforces the substantial nutritional burden carried by children with congenital heart disease in resource limited settings. Chronic hemodynamic stress increases resting energy expenditure and frequently disrupts effective feeding patterns. Pulmonary overcirculation, recurrent respiratory symptoms, and heart failure further increase metabolic demand while reducing caloric intake efficiency. Recurrent illness and prolonged inflammatory stress may also impair nutrient absorption and weight gain during critical periods of childhood growth.[3,4,8,12]

Nutritional impairment appeared across both cyanotic and non cyanotic lesions. Although cyanotic congenital heart disease has traditionally been associated with severe malnutrition because of chronic hypoxemia, ventricular septal defect demonstrated similarly high rates of nutritional problems within this cohort. Large left to right shunts may produce persistent pulmonary congestion, tachypnea, feeding intolerance, and recurrent respiratory tract infections that collectively impair growth despite preserved systemic oxygenation. This observation suggests that chronic hemodynamic burden may contribute to malnutrition as strongly as cyanotic physiology itself.[13,14]

Pulmonary hypertension represented the most common complication identified in the study population. Persistent exposure to increased pulmonary blood flow in untreated or hemodynamically significant shunt lesions likely contributed to this finding. Pulmonary vascular remodeling may develop progressively when left to right shunting remains uncontrolled over prolonged periods. Previous studies have also documented pulmonary hypertension as a frequent complication among pediatric patients with congenital heart disease, especially in settings where delayed diagnosis or limited access to definitive intervention remains common.[3,6,12]

Healthcare utilization patterns demonstrated wide variability across the cohort. Most patients attended outpatient services only once or twice during the study year, whereas a smaller subgroup required repeated follow up with substantially higher visit frequencies. Patients with severe underweight status showed broader healthcare utilization distributions compared with children who maintained normal nutritional status. This pattern may indicate that malnutrition coexists with more severe symptoms, recurrent clinical instability, or persistent feeding difficulties requiring closer monitoring. Higher outpatient visit frequency may therefore reflect cumulative disease burden rather than routine surveillance alone.

Multivariable regression analysis identified young toddlers and toddlers as the age groups with the highest odds of malnutrition. Early childhood represents a vulnerable developmental period characterized by rapid growth velocity and increased nutritional demand. Children with congenital heart disease may struggle to meet these metabolic requirements because chronic cardiac dysfunction limits effective energy utilization and feeding capacity. Transition from exclusive milk feeding toward more varied dietary intake may also contribute to nutritional instability in younger children with persistent respiratory symptoms or fatigue during feeding.[4,8,12]

Higher burden scores independently increased the odds of malnutrition in the final regression model. This finding supports the concept that nutritional deterioration in congenital heart disease reflects cumulative clinical stress rather than a single isolated manifestation. Recurrent respiratory symptoms, feeding problems, pulmonary hypertension, and repeated healthcare encounters may interact synergistically and progressively worsen nutritional reserve. Burden based assessment may therefore provide a broader representation of disease complexity among pediatric congenital heart disease outpatients.

Cyanotic physiology did not demonstrate a statistically significant association with malnutrition after adjustment for covariates. Several explanations may account for this finding. First, the relatively small number of cyanotic cases reduced statistical power for subgroup comparison. Second, non cyanotic lesions with substantial left to right shunting may produce severe metabolic stress and feeding dysfunction comparable to cyanotic lesions. Third, outpatient survivors with cyanotic congenital heart disease may already represent a relatively stable subgroup receiving long term specialized care. Similar observations have been reported by studies demonstrating that nutritional impairment in congenital heart disease depends not only on oxygen saturation status but also on hemodynamic severity and chronic symptom burden.

The present study has several limitations. The retrospective design relied on medical record documentation that demonstrated substantial missingness within symptom and complication variables. Detailed completeness assessment and proportions of missing data across study variables are summarized in S1 Table. Echocardiographic severity parameters, surgical history, socioeconomic conditions, and detailed dietary assessments were not consistently available for analysis. The study also originated from a single tertiary referral center, which may limit generalizability to broader community populations. Despite these limitations, the study included a relatively large outpatient cohort and integrated nutritional assessment, healthcare utilization analysis, and multivariable modeling within a pediatric congenital heart disease population from a resource limited setting.

The findings highlight the importance of early nutritional surveillance and integrated long term outpatient management among children with congenital heart disease. Younger children with recurrent symptoms and higher cumulative clinical burden may require more intensive nutritional monitoring and multidisciplinary follow up to reduce complications and improve long term growth outcomes.

## CONCLUSION

Pediatric outpatients with congenital heart disease at Adam Malik General Hospital were predominantly affected by non cyanotic lesions, with ventricular septal defect, patent ductus arteriosus, and atrial septal defect representing the most common diagnoses. Malnutrition affected nearly half of the study population and remained closely associated with younger age groups, higher cumulative clinical burden, and increased healthcare utilization. Pulmonary hypertension emerged as the most frequently documented complication, reflecting the substantial hemodynamic burden carried by patients with persistent shunt lesions.

Young toddlers and toddlers demonstrated the highest odds of malnutrition, indicating that early childhood represents a vulnerable period for nutritional deterioration among children with congenital heart disease. Increased outpatient visit frequency among malnourished patients also suggested greater long term disease complexity and clinical monitoring needs. These findings emphasize the importance of integrated nutritional surveillance and multidisciplinary outpatient management to improve growth outcomes and reduce chronic disease burden among pediatric congenital heart disease patients in resource limited settings.

## Acknowledgments

The authors would like to thank the Department of Pediatrics and the medical record staff at Adam Malik General Hospital for their assistance during data retrieval and administrative support throughout the study process. The authors also acknowledge the support provided by Faculty of Medicine, Universitas Sumatera Utara in facilitating the conduct of this research.

## Funding

The authors received no specific funding for this work.

## Data Availability Statement

The data underlying the findings of this study contain potentially identifiable patient information derived from retrospective medical records and cannot be shared publicly because of institutional and ethical restrictions. Deidentified data may be made available from the corresponding author upon reasonable request and with permission from Adam Malik General Hospital and the Health Research Ethics Committee of the Faculty of Medicine, Universitas Sumatera Utara.

## Supporting information

**S1 Table. Completeness assessment of variables included in the retrospective pediatric congenital heart disease cohort.** Abbreviations: CHD, congenital heart disease. Data completeness was evaluated across all study variables included in the retrospective outpatient cohort. Symptom- and complication-related variables demonstrated substantial missingness because of inconsistent retrospective clinical documentation. No imputation procedures were performed, and complete-case analysis was applied for multivariable regression modeling.

**S2 Table. Univariable logistic regression analysis of factors associated with malnutrition among pediatric congenital heart disease outpatients.** Odds ratios (ORs) greater than 1 indicate increased odds of malnutrition.

**S3 Table. Sensitivity analysis using the original age-group classification in multivariable logistic regression.** Sensitivity analysis was performed using the original age-group classification to evaluate the robustness of the primary multivariable logistic regression findings. Adjusted odds ratios (ORs) with 95% confidence intervals (CIs) were derived from logistic regression models with malnutrition as the dependent variable. Reference categories were non-cyanotic CHD, female sex, and preschool-aged children. Odds ratios greater than 1 indicate increased odds of malnutrition. *NE = not estimable with reliable precision because of sparse observations and quasi-complete separation in the neonatal subgroup (n = 4). Despite instability in this subgroup, the overall direction and magnitude of associations remained consistent with the primary analysis, supporting the robustness of the primary findings.

**S4 Table. Multicollinearity diagnostics for the multivariable logistic regression model.** *GVIF = generalized variance inflation factor. Adjusted GVIF was calculated as GVIF^(1/(2×Df))^ to account for variables with multiple degrees of freedom. Higher adjusted GVIF values indicate greater collinearity among predictors. All adjusted GVIF values remained below 5, suggesting no evidence of clinically significant multicollinearity within the final multivariable regression model. Df denotes degrees of freedom for each variable.

**S5 Table. Overall performance metrics of the multivariable logistic regression model.** Model performance metrics were calculated to evaluate overall goodness-of-fit, explanatory performance, and classification accuracy of the final multivariable logistic regression model. Lower AIC, AICc, BIC, RMSE, and log-loss values indicate improved model fit. Tjur’s R² reflects explanatory performance for binary outcomes, whereas PCP represents the proportion of correctly classified observations. Collectively, these metrics suggested acceptable overall model performance for exploratory clinical association analysis.

**S1 Fig. Calibration plot of the final multivariable logistic regression model for malnutrition among pediatric congenital heart disease outpatients.** Calibration plot demonstrating agreement between predicted and observed probabilities of malnutrition derived from the multivariable logistic regression model. The dashed diagonal line represents perfect calibration, indicating exact agreement between predicted and observed risk estimates. Blue points and the solid calibration curve represent bootstrap-corrected calibration estimates generated using 1,000 bootstrap resamples. Overall, the model demonstrated acceptable calibration across the observed range of predicted probabilities, although minor deviation from perfect calibration was observed at higher predicted probabilities.

## Notes

### Competing Interest Statement

The authors have declared no competing interest.

### Funding Statement

The author(s) received no specific funding for this work.

### Author Declarations

This study was approved by the Health Research Ethics Committee of the Faculty of Medicine, Universitas Sumatera Utara (Approval number: 653/KEPK/USU/2025). Permission to access medical records was obtained from Adam Malik General Hospital prior to data collection. The study used de-identified retrospective data, and all analyses were conducted in accordance with institutional ethical guidelines.

## REFERENCES

1. Liu Y, Chen S, Zühlke L, Black GC, Choy MK, Li N, Keavney BD. Global birth prevalence of congenital heart defects 1970–2017: updated systematic review and meta-analysis of 260 studies. Int J Epidemiol. 2019 Apr 1;48(2):455–63. doi:10.1093/ije/dyz009. PMID: 30783674; PMCID: PMC6469300.

2. Zhao QM, Liu F, Wu L, Ma XJ, Niu C, Huang GY. Prevalence of congenital heart disease at live birth in China. J Pediatr. 2019 Jan;204:53–8. doi:10.1016/j.jpeds.2018.08.040. Epub 2018 Sep 27. PMID:30270157.

3. Lee J, Marshall T, Buck H, Mulder P, Daack-Hirsch S. Growth failure in children with congenital heart disease. Children (Basel). 2025 May 9;12(5):616. doi:10.3390/children12050616.

4. Khosroshahi AJ, Shoaran M, Ghaffari S, Shabanpour E, Seraj Ebrahimi P, Ansari A, Khosravi R, Azarm E, Sadeghvand S, Erabi G, Chichagi F. Growth pattern of children with congenital heart disease before and after open heart surgery. Front Pediatr. 2025 Jul 24;13:1463998. doi:10.3389/fped.2025.1463998. eCollection 2025. PMID:40777158; PMCID:PMC12328431.

5. Jung SY. Growth in children with congenital heart disease: filling the gap with nationwide evidence. Korean Circ J. 2025 May 27;55(10):936–7. doi:10.4070/kcj.2025.0139. PMID:40537436; PMCID:PMC12599446.

6. Cha JH, Choi YJ, Ryu S, Cho Y, Yang S, Na JY. Physical growth trajectories in children with congenital heart disease: a nationwide study. Korean Circ J. 2025 Oct;55(10):923–35. doi:10.4070/kcj.2025.0020. Epub 2025 Apr 23. PMID:40537424; PMCID:PMC12599410.

7. Benderly M, Buber J, Kalter-Leibovici O, Blieden L, Dadashev A, Lorber A, Nir A, Yalonetsky S, Chodick G, Weitzman D, Balicer R, Dray EM, Murad H, Razon Y, Hirsch R; Israeli Adult Congenital Heart Disease Research Group. Health service utilization patterns among adults with congenital heart disease: a population-based study. J Am Heart Assoc. 2021 Jan 19;10(2):e018037. doi:10.1161/JAHA.120.018037. Epub 2021 Jan 12. PMID:33432841; PMCID:PMC7955316.

8. Abbas Q, Ali H, Ahuja AK, et al. Preoperative nutrition status in children with congenital heart disease and its impact on postoperative outcomes: a systematic review and meta-analysis. Sci Rep. 2025 Jul 16;15:25738. doi:10.1038/s41598-025-96374-z.

9. Thomford NE, Biney RP, Okai E, et al. Clinical spectrum of congenital heart defects detected at the child health clinic in a tertiary health facility in Ghana: a retrospective analysis. J Congenit Heart Dis. 2020 Aug 3;4:3. doi:10.1186/s40949-020-00034-y.

10. Finariawan F, Mahmud SA. The characteristics and distribution of congenital heart disease in outpatient clinic and inpatient ward of RSUD Dr. Soedono Madiun East Java in year 2015. Acta Cardiol Indones. 2018 Jul;4(1):9. doi:10.22146/aci.36633.

11. Baksh AK, Waworuntu DS, Umboh A. Profil penyakit jantung bawaan di Bagian Ilmu Kesehatan Anak RSUP Prof. Dr. R. D. Kandou Manado periode September 2022–Agustus 2023. Acta Cardiol Indones. 2023;12(3):55352. doi:10.35790/ecl.v12i3.55352.

12. Woldesenbet R, Murugan R, Mulugeta F, Moges T. Nutritional status and associated factors among children with congenital heart disease in selected governmental hospitals and cardiac center, Addis Ababa, Ethiopia. BMC Pediatr. 2021 Dec 2;21(1):538. doi:10.1186/s12887-021-03023-1. PMID:34856935; PMCID:PMC8638350.

13. Jone PN, Ivy DD, Hauck A, Karamlou T, Truong U, Coleman RD, Sandoval JP, Del Cerro Marín MJ, Eghtesady P, Tillman K, Krishnan US. Pulmonary hypertension in congenital heart disease: a scientific statement from the American Heart Association. Circ Heart Fail. 2023 Jul;16(7):e00080. doi:10.1161/HHF.0000000000000080. Epub 2023 Jun 26. PMID:37357777.

14. Loni R, Ranganath P, Juvekar M, Tikare N, Bidari LH, Karva MM. Clinical profile and outcome of children with congenital heart diseases admitted with acute events in a paediatric tertiary care unit in North Karnataka. Int J Contemp Pediatr. 2021 Jul;8(8). doi:10.18203/2349-3291.ijcp20212757.

15. Kempny A, Diller GP, Dimopoulos K, Alonso-Gonzalez R, Uebing A, Li W, Babu-Narayan S, Swan L, Wort SJ, Gatzoulis MA. Determinants of outpatient clinic attendance amongst adults with congenital heart disease and outcome. Int J Cardiol. 2016 Jan 15;203:245–50. doi:10.1016/j.ijcard.2015.10.081. Epub 2015 Oct 22. PMID:26519677.

